# Agent-based Investigation of the Impact of Low Rates of Influenza on Next Season Influenza Infections

**DOI:** 10.1101/2021.08.18.21262185

**Authors:** Mary G Krauland, David D Galloway, Jonathan M Raviotta, Richard K Zimmerman, Mark S Roberts

## Abstract

**Introduction:** Interventions to curb the spread of SARS-CoV-2 during the 2020-21 influenza season essentially eliminated influenza during that season. Given waning antibody titers over time, future residual population immunity against influenza will be reduced. The implication for the subsequent 2021-22 influenza season is unknown.

**Methods:** We used an agent-based model of influenza implemented in the FRED (Framework for Reconstructing Epidemiological Dynamics) simulation platform to estimate cases and hospitalization over two succeeding influenza seasons. The model uses a synthetic population to represent an actual population, and individual interactions in workplaces, school, households and neighborhoods. The impact of reduced residual immunity was estimated as a consequence of increased protective measures (e.g., social distancing and school closure) in the first season. The impact was contrasted by the level of similarity (cross-immunity) between influenza strains over the seasons.

**Results:** When the second season strains were dissimilar to the first season (have a low level of cross immunity), a low first season has limited impact on second season cases. When a high level of cross-immunity exists between strains in the 2 seasons, the first season has a much greater impact on the second season. In both cases this is modified by the transmissibility of strains in the 2 seasons. In the context of the 2021-22 season, the worst case scenario is a highly transmissible strain causing increased cases and hospitalizations over average influenza seasons, with a possible significant increase in cases in some scenarios. The most likely overall scenario for 2021-22 is a more modest increase in flu cases over an average season.

**Discussion:** Given the light 2020-21 season, we found that a large, compensatory second season might occur in 2021-22, depending on cross-immunity from past infection and transmissibility of strains. Furthermore, we found that enhanced vaccine coverage could reduce this high, compensatory season. Young children may be especially at risk in 2021-22 since very young children were unlikely to have had any exposure to infection and most immunity in that age group would be from vaccination, which wanes quickly.

## Introduction

SARS-CoV-2 was declared a pandemic by the World Health Organization (WHO) on March 11, 2020. Cases were identified in the US in early 2020, resulting in widespread interventions to reduce spread of the virus, including school closure and measures to decrease social interactions. These interventions impacted not only COVID-19 transmission but also transmission of other diseases that spread by the same mechanisms. Little influenza activity was seen in the northern hemisphere during the fall and winter of 2020-21. In Finland, a decreased incidence of respiratory infections was noted when SARS-CoV-2 restrictions were instituted in March 2020[1]. Northern California reported decreased influenza activity once Sars-CoV-2 interventions were enacted[2]. Limited influenza transmission was detected during the summer influenza season in the southern hemisphere[3]. Influenza activity was similarly limited in Canada[4] and in Europe, where the WHO European Region saw a 99.8% reduction in detection of influenza by sentinel surveillance over weeks 40/2020 and 8/2021 relative to the same period in the 6 prior seasons[5]. Little influenza activity was detected by the Centers for Disease Control and Prevention (CDC) in the US during the 2020-21 influenza season[6].

Prior year vaccination is believed to provide little protection to subsequent year influenza, but natural infection appears to provide measurable immunity for several seasons, particularly if the circulating strains are close antigenic matches[7, 8]. However, the limited number of influenza cases in the US in 2020-21 has raised concerns about the possibility of a higher burden of influenza illness in the 2021-22 season due to reduced immunity from prior year natural infection, even if a trend to higher acceptance of influenza vaccination continues. An additional complication is the difficulty in choosing strains for the 2021-22 vaccine because a limited number of surveillance samples were available. Members of the population under the age of 2 may be at increased risk after a season with limited influenza since they likely have never been exposed to natural infection and therefore may be at higher risk.

Agent-based modeling can be used to perform highly detailed investigations of possible disease scenarios. The FRED (Framework for Reconstructing Epidemiologic Dynamics) simulation platform[9] is an agent-based modeling platform that was developed in response to the 2009 influenza pandemic and has been used extensively for modeling influenza as well as other infectious and non-infectious conditions [10-13]. To explore the impact of limited influenza activity on subsequent year influenza burden, we used FRED to model 2 season influenza scenarios under a variety of assumptions on the impact of natural infection on second year immunity and a variable degree of antigenic relatedness of first and second year circulating strains. We performed simulations to estimate the impact of increased vaccination and the specific impact on the youngest age group in the population.

## Methods

FRED is an agent-based modeling platform that facilitates modeling of diverse conditions. The FRED simulation platform has been described in detail previously[9]. Briefly, FRED uses census-based synthetic populations whose members have demographics that are statistically equivalent to real populations at the county level. Agents have explicit geographic household locations, and their household makeup is statistically similar in demographics and income to real populations. Agents interact daily in locations including their household, neighborhood, school and workplace, as appropriate. Infectious conditions spread through the population due to these interactions. Populations are available for all counties of the US and simulations can be performed on aggregations of counties or subpopulations down to the census block group level.

### Influenza Model

The FRED influenza model is a modification of a basic SEIR model containing the following states: Susceptible; Exposed; Pre-symptomatic; Infected symptomatic; Infected asymptomatic; Hospitalized; Recovered; and Died (Figure 1).

**Figure 1.**
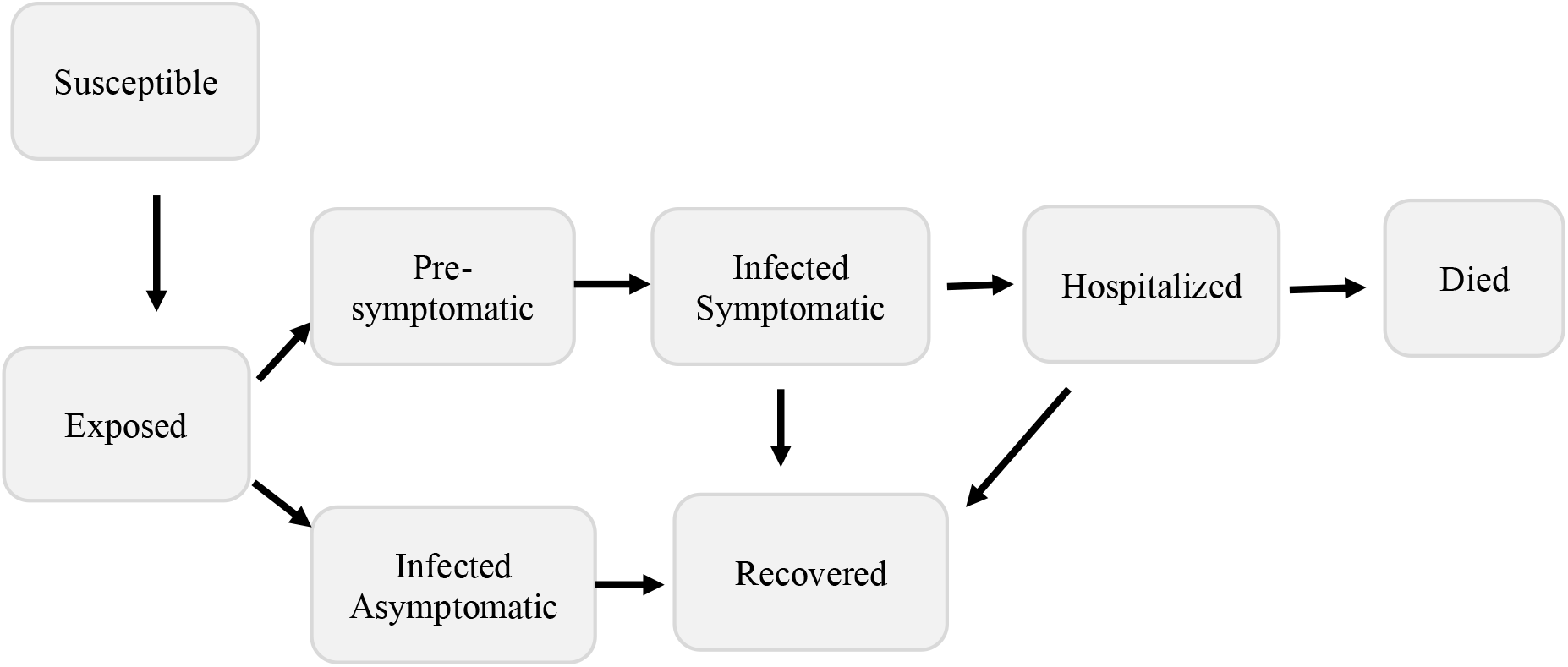
Influenza model diagram.

Simulations were performed on Allegheny County, a medium sized county in the southwest of Pennsylvania, with a population of ∼1.2 million. It contains both urban and suburban areas. Due to its size, Allegheny County is a convenient population for modeling influenza. Simulations start on September 15, 2020. Agents are initialized to the Susceptible state and wait for infection. Outbreaks are started by introducing cases into susceptible agents. In this model 20 new cases are injected on Nov 15 of both simulation years. Further infection is result of probabilistic interaction with infectious agents. Exposed agents become pre-symptomatic (75%) or asymptomatic (25%). Susceptible agents in any state can be infected. This model includes an infectious pre-symptomatic state of length 1 day, with agents in that state being half as infectious as agents in the infected symptomatic state. Pre-symptomatic agents move to the symptomatic infected state. Symptomatic infected agents remain in that state for a number of days drawn from a lognormal distribution with median 4 and dispersion1.5. This model also includes an infectious asymptomatic state with infectivity half that of symptomatic agents. Agents remain in that state for a number of days drawn from a lognormal distribution with median 5 and dispersion 1.5. Asymptomatic infected agents are half as infectious as agents in the infected symptomatic state. Symptomatic agents may move to the Hospitalized state at age-structured rates obtained from CDC data [14]. Agents in the Hospitalized state die at age-appropriate rates. Asymptomatic infected, symptomatic infected and Hospitalized agents who do not die move to the Recovered state. Recovered agents have susceptibility to the infecting influenza strain set to zero; this susceptibility increases at a rate of 3% per month. Cross-immunity to a second season strain is implemented by reducing susceptibility to second strain in agents infected in the first season.

Influenza utilizes a stay-at-home behavior module. Symptomatic agents stay at home and so do not interact in their neighborhood, workplace, or school, with a probability of 50%.

### Vaccination Model Details

Agents are immunized by age group as per CDC data using the following rates: age 0.5-17, probability 0.504; age 18-49 probability 0.342; age 50-64 probability 0.468; age>=65 probability 0.687[15]. In the model, vaccination reduces susceptibility to influenza by 40% (vaccination efficacy in CDC studies ranges between 10-60%)[16, 17].

Strain specific vaccination occurs in October of the simulation year and immunity wanes after vaccination at 7% per month[7].

### Transmissibility Parameter in FRED

FRED does not specify an explicit value for the basic reproduction number (R_0_) or effective reproduction number (R) for a simulated infection. In FRED, an infectious condition has a transmissibility parameter and an infectious period that are characteristic of the simulated disease, and these are set as part of the disease model. The population used for the simulation will have characteristics, such as demographic and geographic makeup and contact patterns, which are drawn from the census-based synthetic populations used for the simulation. The transmissibility, infectious period and population characteristics combine to generate an R or R_0_ in a given simulation. R or R_0_ are therefore not an input to the model but rather are outputs produced by the interaction of a disease model and a specific population. The disease model is typically calibrated to produce an R or R_0_ that is realistic for a modeled disease. We used 2 values for the transmissibility input parameter for simulations, to represent 2 levels of influenza transmissibility (0.6 and 0.8). A transmissibility parameter of 0.6 produces an initial reproduction number of 1.01(SD 0.23) in a model including vaccination. A transmissibility parameter of 0.8 produces an initial reproduction number of 1.39 (SD 0.30) in the same model. The mean influenza reproduction number is estimated to be 1.3 (range 0.9-2.1)[18].

### COVID-19 Protections Model

We used protections developed for a COVID-19 model to investigate impact on influenza. In FRED all schools can be closed on specified dates. In the model, all schools were closed on March 15, 2020, and reopened on April 15, 2021. After reopening, schools operated on a regular summer closure schedule. When schools are closed, agents do not interact in school or classroom, including agents assigned as teachers. In the model, social distancing is accomplished by selecting agents to isolate in their homes, thereby removing neighborhood, school and workplace interactions for those agents. Social distancing was set at 60% of the population as indicated by mobility data collected during the COVID-19 pandemic. In 2 year simulations using the COVID-19 protections model, it was applied only in first season.

### Increased Influenza Vaccination due to COVID-19 vaccination spill-over

To estimate the impact of increased vaccination, selected simulations were run with 10% and 20% increased vaccination levels. Influenza vaccination levels as reported by the CDC from 2010 to 2020 have increased by levels in the 10-20% range[19].

### Impact on age 0-4

Selected simulations were run with the addition of actuarial maternity and mortality models to add agents by birth to the population during the simulation. The model allowed prediction of cases and hospitalizations for agents age 0-4 during the simulations.

## Results

### Base Model Results

Without vaccination the low transmission scenario (reproduction number 1.01) produced an attack rate of 12.7% and the high transmission scenario (reproduction number 1.39) produced an attack rate of 30.3%. Vaccination according to the coverage rates reported by the CDC reduced the attack rate of the low transmission scenario to 4.4% and the high transmission scenario attack rate to 23.3%.

### Impact of School Closure and Social Distancing on Influenza Model

To check the model’s integrity, we simulated 2 seasons with identical strains with 100% antigenic similarity and 3% waning of immunity per month. In this scenario, residual first season immunity prevented any influenza outbreak in second season. This integrity check was passed.

In a second integrity check, when schools were closed and social distancing at 60% was added to the model, the first-year season was completely prevented and the second season produced case counts similar to the first season, showing complete removal of the residual immunity effect.

In more realistic situations, when varying levels of social distancing but no school closure were applied in the model, increased levels of social distancing in season 1 caused decreased season 1 influenza cases, resulting in increased season 2 cases (Figure 2). This has significance for public health. In this scenario, 40% social distancing was sufficient to prevent any first season cases. In the 2020-21 influenza season, Allegheny County surveillance reported 328 lab confirmed influenza cases compared to 13,889 in 2019-20[20]. In the model, 40% social distancing produced a median of 965 cases during that season (724 symptomatic cases). Since the surveillance count would underestimate total cases, due to under ascertainment, the numbers are roughly comparable between the model results and cases identified by surveillance.

**Figure 2.**
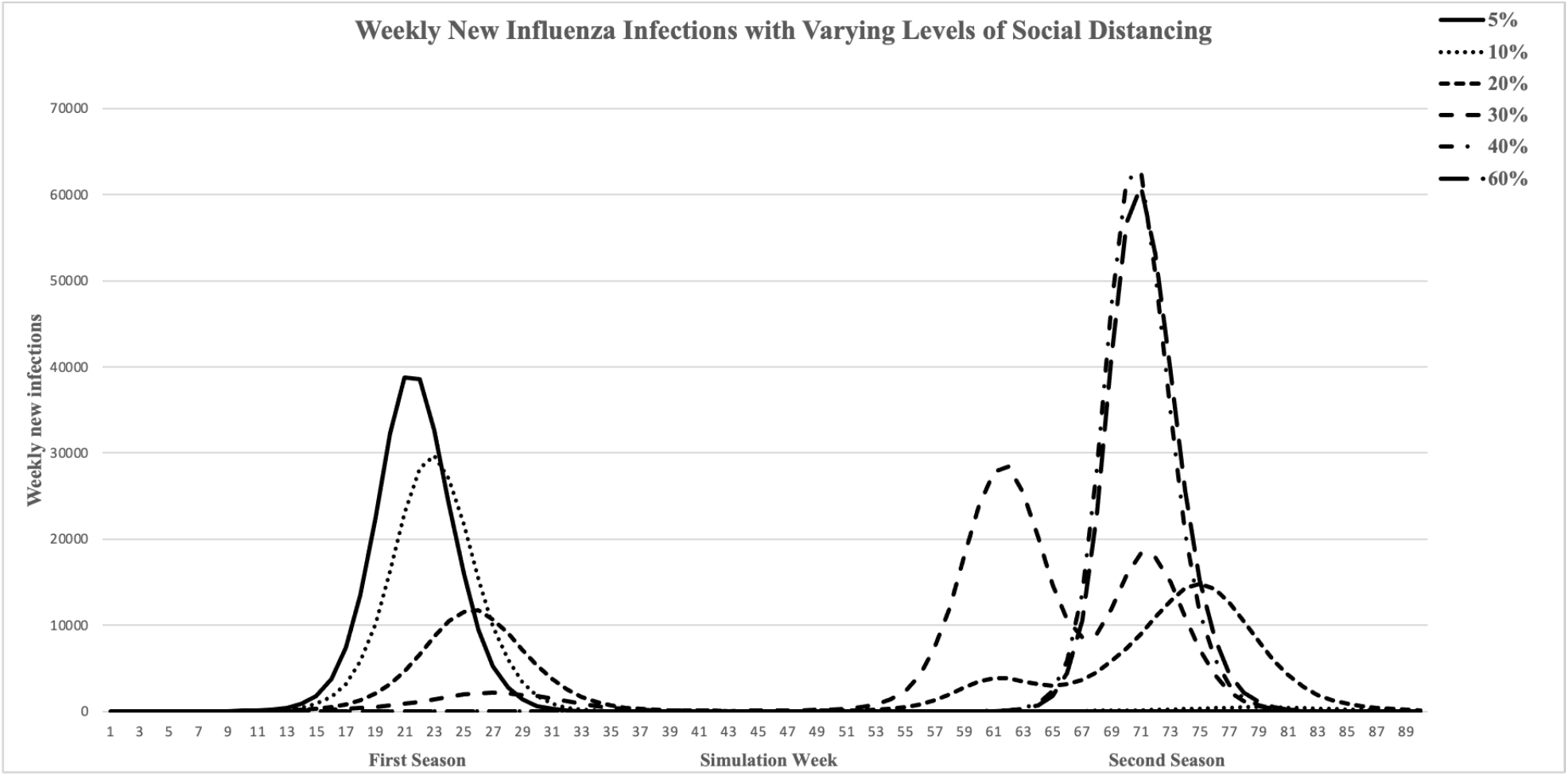
Influenza model incidence curves. Increased levels of social distancing (but no school closure) resulted in decreased first season cases and increased second season cases. Social distancing at 40% prevents seasonal influenza outbreak. Social distancing at 20% and 30% produces highly stochastic second season outbreaks

In some scenarios, season 2 became highly stochastic, such that if the mean of multiple simulations was plotted, the typical curve became distorted because second year outbreaks were not synchronous even though started at the same date.

### Impact of strain transmissibility and cross-immunity on second season influenza cases

In a 2 strain, 2 season model, second season influenza cases and hospitalizations prevented by first year infections varied by the transmissibility of the first and second year strains and by the similarity (cross-immunity) between those strains (Table 1). Decreases in cases and hospitalizations produce similar but not identical results. When both season strains had lower transmissibility, the impact of first season on second season was less pronounced, presumably because the total cases in the first season were lower, with maximum reduction in second season cases of 75.87% and reduction in hospitalizations of 75.76% (Figure 3A). A lower transmissibility strain in the first season had much lower impact on second season cases when the second season strain was more transmissible, only causing a 55.33% reduction in cases and 57.37% reduction in hospitalizations with complete cross-immunity (Figure 3B). When the first season strain had a greater transmissibility than the second season strain, almost all cases and hospitalizations were prevented by even 40% cross-immunity (Figure 3C). When the first season strain was more transmissible, cases and hospitalizations in the second season were eliminated for strains with the same higher transmissibility when cross-immunity was 60% or greater (Figure 3D).

**Table 1.** Percent decrease in influenza cases and hospitalizations compared to baseline by percent cross-immunity between first and second season influenza strains.

|  | low/low<sup>1</sup> | low/high | high/low | high/high |
| --- | --- | --- | --- | --- |
| 10 | 12.94 (14.46) | 1.68(2.20) | 42.51(44.79) | 6.45(8.02) |
| 20 | 24.07 (24.99) | 2.97(3.70) | 74.78(75.78) | 17.39(20.46) |
| 30 | 32.71(32.81) | 4.91(5.57) | 90.78(91.26) | 30.67(35.41) |
| 40 | 42.62 (43.22) | 8.60(10.15) | 96.74(96.63) | 45.80(51.14) |
| 50 | 55.65 (55.15) | 14.43(16.70) | 98.85(98.67) | 76.65(80.20) |
| 60 | 64.55(64.78) | 21.19(23.92) | 99.15(99.01) | 96.21(96.69) |
| 70 | 69.06 (68.90) | 30.14(33.34) | 99.22(99.10) | 98.84(98.79) |
| 80 | 71.43 (71.28) | 38.84(41.92) | 99.23(99.13) | 98.97(98.90) |
| 90 | 74.56 (74.48) | 45.61(48.08) | 99.24(99.13) | 98.99(98.92) |
| 100 | 75.87 (75.76) | 55.33(57.37) | 99.21(99.13) | 98.98(98.94) |

<sup>1</sup> Transmissibility first season / transmissibility second season as defined in Methods
<sup>2</sup> Cases (Hospitalizations)

**Figure 3.**
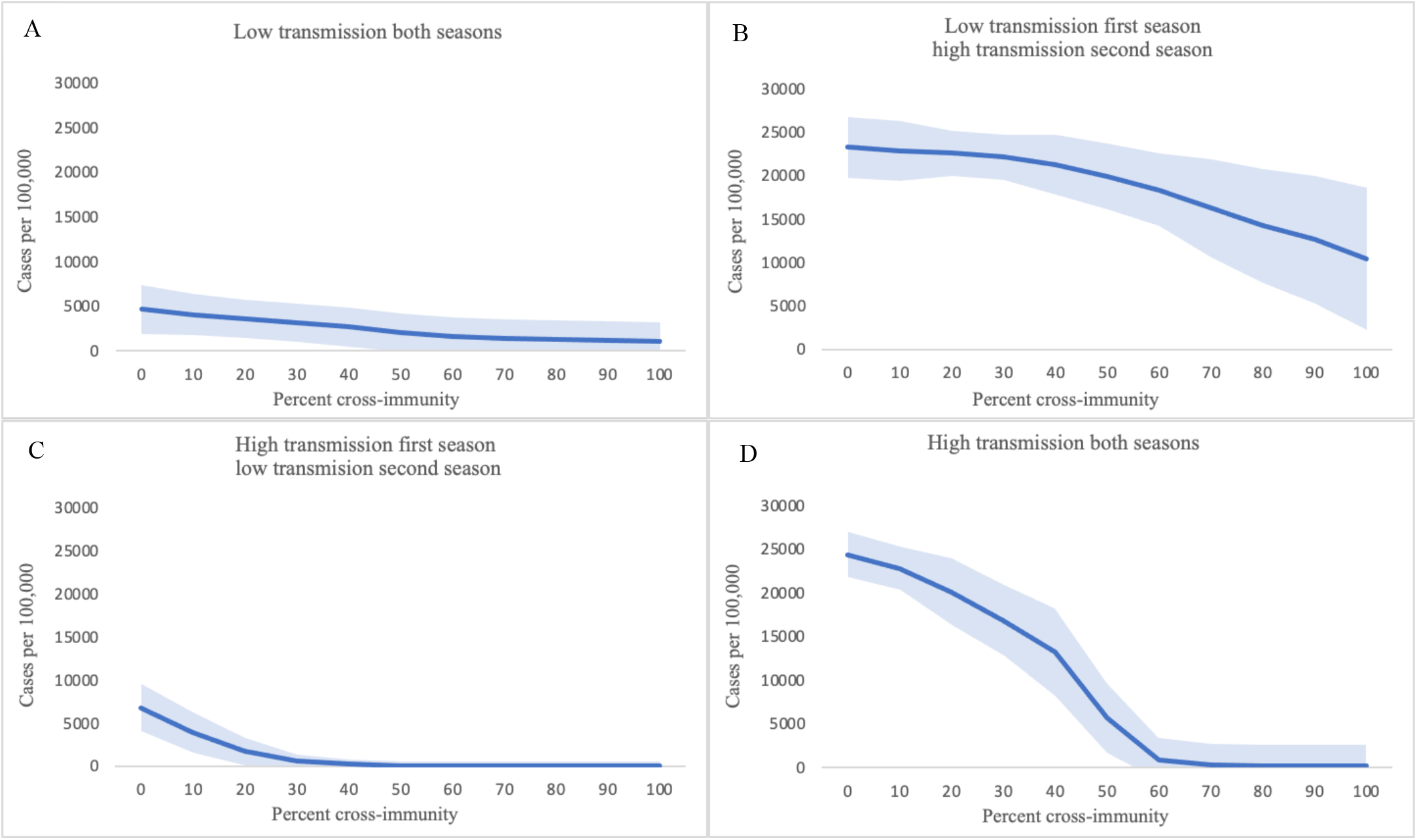
Influence of cross-immunity between first and second season strains on the impact of first season influenza cases on second season influenza cases. X-axis, % cross-immunity between first- and second-year strains; Y-axis, mean second season cases per 100,000. Shaded area represents +/- one standard deviation from mean.

### Impact of Increased Vaccination

Increasing vaccination rates above those reported by the CDC by 10% and 20% decreased the number of cases and hospitalizations during a single season. Increased vaccination rates had a greater impact on simulations with a lower transmissibility strain (10% increase in vaccination caused a 35.7% decrease in cases and 45.5% decrease in hospitalizations; 20% increase in vaccination caused a 39.7% decrease in cases and 49.7% decrease in hospitalizations). For simulations with a more transmissible strain, increased vaccination resulted in a much more modest effect (10% increase in vaccination caused a 4.4% decrease in cases and 6.5% decrease in hospitalizations; 20% increase in vaccination caused a 7.1% decrease in cases and 10.8% decrease in hospitalizations). In the low transmissibility scenario, increased vaccination rates may have greater impact due to vaccination levels approaching what would be expected to achieve herd immunity due to low reproduction number in that model.

### Impact of Increased Vaccination Coverage on Infants and Young Children

The age group 0-4 makes up 5.17% of the simulation population and accounts for ∼6% of cases in the simulation (6.0 to 6.5% of cases). Increasing vaccination coverage by 10% decreased cases in this age group by 35.7% in the lower transmissibility scenario but only 3.9% in a higher transmissibility scenario (Figure 4). A 20% increase in vaccination coverage resulted in 40.4% fewer cases for a lower transmissibility strain but only 6.4% fewer for a more transmissible strain. Hospitalizations decreased by similar amounts (Table 1; for low transmissibility scenario, 37.1% decrease with 10% coverage increase and 39.1% with 20% increase; for high transmissibility scenario 3.7% decrease with 10% coverage increase and 6.7% with 20% increase).

**Figure 4.**
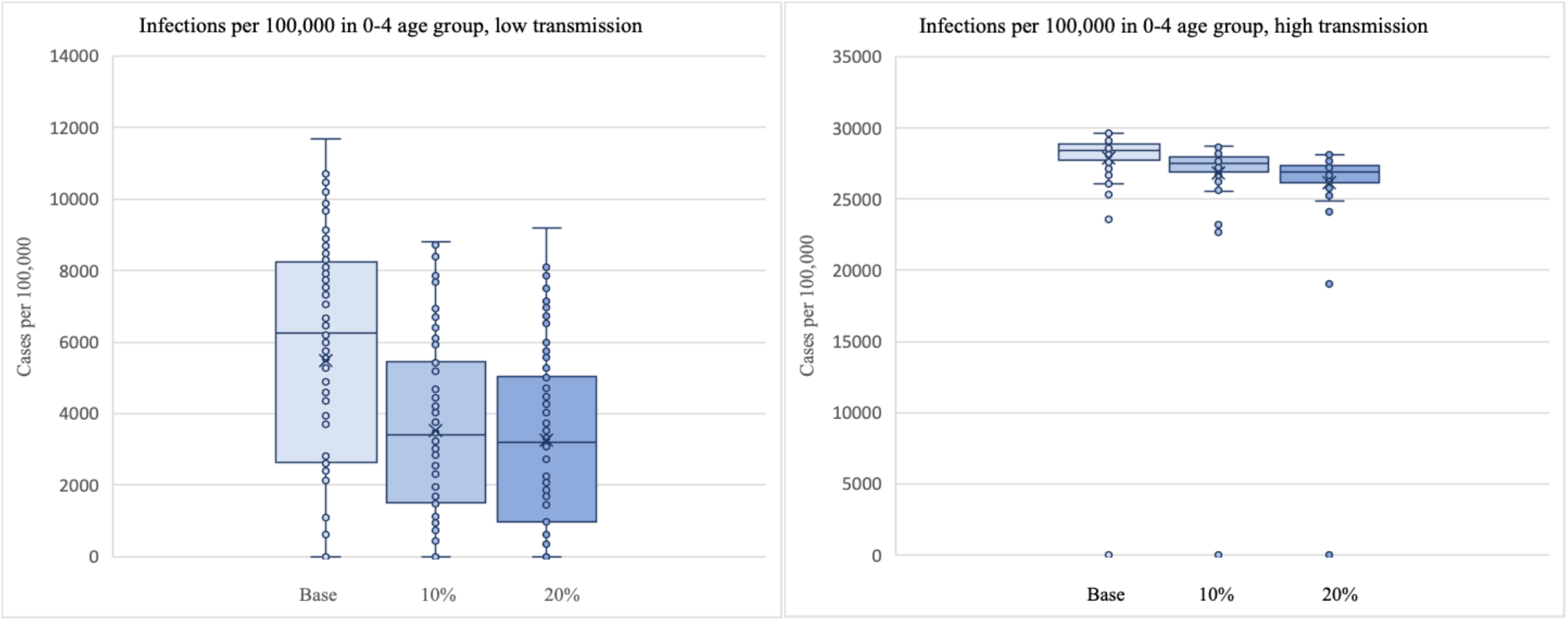
Change in total infections per 100,000 children age 0-4 with increase in vaccination coverage rate. Base vaccination from reported rates. Coverage rates were increased by 10% and by 20%.

### Implication for second season influenza

A possible range of increase in second season cases from low first season rates can be estimated from the modeled second season impact of varied case rates in the first season. This impact in our model is influenced by the transmissibility of strains in the first and second seasons and by antigenic similarity of the two strains. Likely ranges for highly antigenically related strains might be to have 60-80% cross-immunity and less related strains might provide 10-30% cross-immunity. It has been estimated that >80% cross-immunity with a prior outbreak strain is sufficient to prevent transmission of a later strain[21, 22]. The model suggests that in a high transmission scenario (strain ∼R_0_ 1.39) ∼99% of cases and hospitalizations would be prevented in a second season if strains were closely related (Table 1). This level of residual immunity would prevent a strain from circulating. When first and second season strains are less related in a higher transmission scenario, the increase in expected cases would be ∼7-30% increase in infections due to lower immunity. In a low transmission scenario (strain ∼R_0_ 1.01) with closely related strains >65% of cases would be prevented while less related strains would reduce cases by ∼10-30%.

## Discussion

We found that a light first season (such as 2020-21) could result in a large, compensatory second season (such as might occur in 2021-22). This increase could be as large as a doubling of cases, although this is an unlikely scenario. Furthermore, we found that enhanced vaccine coverage could reduce but not eliminate this high, compensatory season.

Influenza strains are highly variable, with genetic drift antigens contributing to emergence of new variants that escape immunity and enable yearly seasonal outbreaks despite population immunity and vaccination. The yearly burden of symptomatic influenza A estimated by the CDC varied by over 4-fold in the past 20 years[23]. Years in which the subtype of the predominant circulating strain changed were not consistently correlated with increases in infection [24]. This underscores the complex nature of influenza immunity and the difficulty in predicting future seasonal outbreaks.

The northern hemisphere 2020-21 season was an anomaly, lacking the usual seasonal outbreak. This was likely due to COVID-19 interventions, such as masking, school closure, remote work and other social distancing interventions. While lack of an influenza outbreak in that season was beneficial in that health care systems were already strained by the COVID-19 pandemic, there is concern that the lack of a 2020-21 influenza season will cause a more severe influenza season in 2021-2022, due to lack of residual immunity resulting from prior season infection. While vaccination is an effective tool to prevent influenza, vaccination rates are typically sub-optimal, and vaccination does not appear to confer long lasting immunity. In response, we modeled this situation.

To help in understanding the impact of an influenza season with very low cases on cases and hospitalizations in the succeeding season, we used an agent-based model of influenza with varying levels of cross-immunity between seasonal strains. We applied school closure and decreased social interactions to agents in the simulation to mimic the interventions enacted to combat COVID-19. In our model, these interventions resulted in a nearly complete elimination of influenza when the interventions were in place. When a limited influenza season was followed by a second influenza season, cases rebounded to a level expected with no population immunity.

We modeled 2 levels of transmissibility of strains and varying levels of cross-immunity between strains, to account for situations in which the highly related and less related strains circulate in successive seasons. In our results, with a low level of cross-immunity, corresponding to poor match between strains, a low first season has limited impact on second season cases. This situation might be considered comparable to an H3N2 predominant season after an H1N1 season. If there is a good antigenic match between seasons and, therefore, a high level of cross-immunity, the model predicts a large impact on second season cases from first season infections. We modeled the cross-immunity resulting from antigenic similarity between first and second season cases from 0 to 100%. It is unlikely that identical strains would predominate in 2 successive seasons (100% cross-immunity scenario); this much immunity would prevent a strain from becoming dominant. Even strains from different subtypes are believed to share some conserved epitopes, so the 0% cross-immunity scenario is also unlikely[25].

Strains with higher levels of transmissibility cause more cases and therefore have a larger impact on second season cases. When the first season has a less transmissible strain, even high levels of cross-immunity prevent fewer cases in the second season. A more transmissible strain in the first season can almost completely prevent cases by a closely antigenically related but less transmissible strain in the second season.

We investigated the effect of increasing vaccination rates by 10 or 20%. This increase could help offset the reduced population immunity resulting from the lack of influenza in 2020-21. This could be particularly important for young children, who may be especially at risk in 2021-22. Very few very young children had influenza in 2020-21 so essentially all immunity in that age group would be from vaccination, which wanes relatively quickly.

Several factors suggest that, while an increase in cases may be expected after a very low first season, the increase in second season cases will not reach what would be expected for a pandemic strain. The most prevalent influenza subtype often switches from one season to next, for example H1N1 predominated in the 2013-14 season while H3N2 predominated in the 2 preceding seasons and these influenza subtype switches, in which cross-immunity would be expected to be limited, do not always cause increased intensity[26]. Immunity due to infection is believed to wane over 3-8 years for H1N1 and 3-5 for H3N2[8] so substantial immunity can be expected to remain from prior seasons. Additionally, residual immunity from exposure to infection early in life is believed to confer life-time immunity to the infecting strain, possibly including some immunity to conserved epitopes[27].

## Strengths and Limitations

FRED is a well-established platform for one season of influenza that we have expanded to a multi-year platform; this was the first publication of that endeavor. The 2021-22 influenza season may be unique in that it lacks any significant 2020-21 impact of residual disease immunity. We have examined it for face validity using influenza infectious disease experts, explored the impact of various parameters and averaged multiple simulations. However, all models are attempts to predict reality, which cannot be known until the season has passed.

## Conclusions

Given the light 2020-21 season, we found that a large, compensatory second season might occur in 2021-22, depending on cross-immunity from past infection and transmissibility of strains. Furthermore, we found that enhanced vaccine coverage could reduce this high, compensatory season. Achieving high vaccination levels in very young may be particularly important in 2021-22, due to lack of immunity from prior infection in that age group.

## Data Availability

All data referred to in the manuscript is publicly available.

## Acknowledgements

The authors would like to acknowledge Donald S. Burke, MD for valuable contributions to the development of this project and John J Grefenstette, PhD for development of the FRED model of social distancing used in some simulations.

